# Auditory grouping ability predicts speech-in-noise performance in cochlear implants

**DOI:** 10.1101/2022.05.30.22275790

**Authors:** Inyong Choi, Phillip E. Gander, Joel I. Berger, Jean Hong, Sarah Colby, Bob McMurray, Timothy D. Griffiths

## Abstract

**Objectives:** Cochlear implant (CI) users exhibit a large variance in understanding speech in noise (SiN). Past works in CI users found that spectral and temporal resolutions correlate with the SiN ability, but a large portion of variance has been remaining unexplained. Our group’s recent work on normal-hearing listeners showed that the ability of grouping temporally coherent tones in a complex auditory scene predicts SiN ability, highlighting a central mechanism of auditory scene analysis that contributes to SiN. The current study examined whether the auditory grouping ability contributes to SiN understanding in CI users as well.

**Design:** 47 post-lingually deafened CI users performed multiple tasks including sentence-in-noise understanding, spectral ripple discrimination, temporal modulation detection, and stochastic figure-ground task in which listeners detect temporally coherent tone pips in the cloud of many tone pips that rise at random times at random frequencies. Accuracies from the latter three tasks were used as predictor variables while the sentence-in-noise performance was used as the dependent variable in a multiple linear regression analysis.

**Results:** No co-linearity was found between any predictor variables. All the three predictors exhibited significant contribution in the multiple linear regression model, indicating that the ability to detect temporal coherence in a complex auditory scene explains a further amount of variance in CI users’ SiN performance that was not explained by spectral and temporal resolution.

**Conclusions:** This result indicates that the across-frequency comparison builds an important auditory cognitive mechanism in CI users’ SiN understanding. Clinically, this result proposes a novel paradigm to reveal a source of SiN difficulty in CI users and a potential rehabilitative strategy.

## Introduction

Although cochlear implants (CIs) have been the most successful intervention for patients with severe sensorineural hearing loss, persistent variability in CI speech-perception outcomes remains (Gantz et al., 2016). Although much of this variability derives from peripheral factors such as the degree of current spread that affects spectral resolution (Bingabr et al., 2008) and/or changes to the spectral mapping in the periphery (Hamzavi et al., 2003), established CI users with similar audiometric profiles and spectral resolution still differ when confronted with noise (Fetterman & Domico, 2002; Noble et al., 2009). This suggests that perceptual and cognitive processes may account for differences in speech perception performance. However, the neural and computational mechanisms that underlie these central processes are poorly understood.

Studies in normal auditory systems have considered speech-in-noise (SiN) understanding as a “cocktail party problem” (Cherry, 1953), a problem of extracting a target sound from intermixed competing sounds. A solution for the cocktail party problem is a successful auditory scene analysis (Bregman, 1994), which can be understood as a chain of processes including 1) sensory encoding of acoustic dynamics, 2) grouping of acoustics features to form auditory objects (Darwin, 1997), and 3) across-object competition. Under this ASA framework, individual differences in SiN understanding may originate from each of the ASA processes. Evidence exists of individual differences in such ASA functions including encoding of suprathreshold dynamics (Ruggles et al., 2011) and auditory grouping (Holmes & Griffiths, 2019; Holmes et al., 2021).

*In cochlear implant users*, most previous works investigating the sources of outcome variance have focused on the status of the peripheral auditory system, that is only related to the first process above (i.e. encoding acoustic cues) (Winn et al., 2016; Won et al., 2007), leaving a gap in our understanding of how the central auditory functions affect speech-in-noise perception in CI users.

Our premise is that auditory cognitive processes, independently from peripheral processing, also contribute to speech-in-noise understanding in CI users. Especially, auditory object formation is a key process for target identification in crowded auditory scenes. One of the most effective perceptual strategies for object formation is grouping frequency components with temporal coherence (Shamma et al., 2011; Teki et al., 2016; Teki et al., 2013; Teki et al., 2011). Because most CI programming schemes deliver relatively intact temporal envelope patterns, theoretically, temporal coherence can be perceived by electric hearing.

This study aimed to promote our understanding of a relationship between the central auditory functions and speech-in-noise outcomes in CI users by testing whether an auditory figure-ground task, an established paradigm that tests temporal coherence detection ability, explains the further amount of variance in CI outcomes in addition to spectral and temporal resolution. A relatively large number (47) of post-lingually deafened CI users were recruited for this study and performed the stochastic figure-ground task (SFG: (Teki et al., 2011)) in which listeners detect temporally coherent tone pips in the cloud of many tone pips that rise at random times at random frequencies. The same subjects also performed spectral ripple discrimination and a temporal modulation detection task as well as a sentence-in-noise understanding task (AzBio: (Spahr et al., 2012)). The accuracy of SFG, spectral ripple discrimination, and temporal modulation detection tasks were used as three predictor variables of a multiple linear regression model to predict the AzBio performance, to test the hypothesis that the performance on the SFG task explains a further amount of variance in CI users’ AzBio performance that is not explained by spectral and temporal resolution.

## Materials and Methods

### Participants

Forty-seven CI users, between 20 and 79 years of age (mean = 60.9 years, SD = 12.1 years; median = 63.3 years; % female), were recruited from the University of Iowa Cochlear Implant Research Center. All the participants were post-lingually deafened and neurologically normal. The average length of device use was 39.5 months (SD = 56.8 months). The average duration of deafness was 22.0 years (SD = 15.0 years). Five subjects were bilateral CI users. Among the remaining subjects, 66.1% had CI in the right ear. 76.3% were Hybrid CI users (i.e., electric acoustic stimulation within the same ear). Average threshold of low-frequency (i.e., 250 and 500 Hz) residual acoustic hearing was 59.4 dB HL (SD = 20.5 dB HL). American English was the primary language for all the participants. Most participants were tested during the same day as a clinical visit in which they received an annual audiological examination and device tuning. Most of these CI users were bimodal or hybrid CI users who would have some residual acoustic hearing. Duration of device use was obtained from clinical records. All study procedures were reviewed and approved by the local Institutional Review Board.

### Task design and procedures

All CI users performed the spectral ripple discrimination, temporal modulation detection, SFG, and speech in noise (AzBio).

#### Speech-in-noise: AzBio

Performance on a sentence-in-noise task (AzBio; (Spahr et al., 2012)) was used as a dependent variable in the later multiple linear regression analysis to predict CI individuals’ SiN ability. Our AzBio task was performed at +5dB SNR condition at 70dB SPL in a double-walled sound booth using a sound field presentation. A loudspeaker was located 1.5 m in front of the subject. A single loudspeaker was used for both speech and noise (i.e., diotic listening). Subjects had to repeat the sentence they heard. Audiologists counted the number of currently repeated words from the outside of the sound booth. The ratio of currently repeated words to the total number of words in all the presented sentences was determined and used as an individual subject’s accuracy for later analyses.

#### Spectral Ripple and Temporal Modulation: Stimuli

The stimuli for both tasks were generated in MATLAB at the time of testing based on the parameters set by the Updated Maximum-Likelihood (UML) adaptive procedure (Shen et al., 2015). Starting parameters were based on pilot data from 40 CI users, and subsequent trials were adaptively generated based on the predictions of the UML procedure given the subject’s responses. Unlike traditional tasks, the UML procedure adaptively predicts what to test to best estimate an individual’s psychometric function. Estimates of the psychometric function were updated after every trial.

For the spectral ripple task, stimuli were broadband noise with sinusoidal variations (ripples). The ripple peaks were evenly spaced on a log-frequency scale with the density of ripples held constant at 1.25 ripples per octave. The amplitude depth of the ripples (in dB) was manipulated based on the UML predictions. Two standard sounds were created with a randomized starting location for the spectral peak and the oddball was created with an inverted phase to be maximally distinct.

For the temporal modulation detection task, stimuli were a complex tone comprised of component frequencies at 1515, 2350, 3485, 5045, and 6990 Hz. A sine wave was overlaid on the complex tone to modulate its amplitude. The envelope frequency was 20 Hz and the depth of the sinusoid was determined by UML prediction. Trials either had two modulated sounds, where the oddball was unmodulated, or two unmodulated sounds, where the oddball was modulated.

Stimuli for both tasks were 500 ms in duration and ramped with a 50 ms rise/fall. To compensate for intensity differences in the modulated stimuli, root mean square values were equalized and the presentation level was roved randomly, so loudness could not be used as a cue.

#### Spectral Ripple and Temporal Modulation: Procedure

Spectral and temporal resolutions were determined by spectral ripple discrimination and temporal modulation detection tasks using a 3-alternate forced-choice oddball detection paradigm. The task was implemented using Psychtoolbox 3 ((Brainard, 1997); (Pelli, 1997)) in MATLAB (The Mathworks). On each trial, two standard stimuli and one oddball were played in random order with an ISI of 750 ms. A numbered box appeared on the computer screen as each stimulus played. Subjects were instructed to choose the token that differed from the other two. The UML approach allowed the tasks to be much shorter than traditional staircase measures; each task was 70 trials. Both tasks began with 4 practice trials to familiarize the subject with the task and correct/incorrect feedback was given on every trial.

#### SFG: stimuli

The SFG stimuli were generated using the same design principle described in (Teki et al., 2011); figure and ground tone pips overlap in time and frequency space, and they can be distinguished only by their fluctuation statistics. In 50% of the trials, the tonal components repeated in frequency over 2 seconds, which popped out as “figure” out of “ground.”

In our experiment, the frequencies of the figure varied from trial to trial but the number of figure tone pips was always six. To avoid subjects’ spectral resolution or low-frequency residual acoustic hearing confounding their SFG ability, the frequency gap between adjacent figure tone-pips (i.e., repeated tonal components) was at least one octave and they were all above 1kHz. Figure 1 shows an example schematic stimulus containing the “figure” tone pips in the second half.

**Figure 1.**
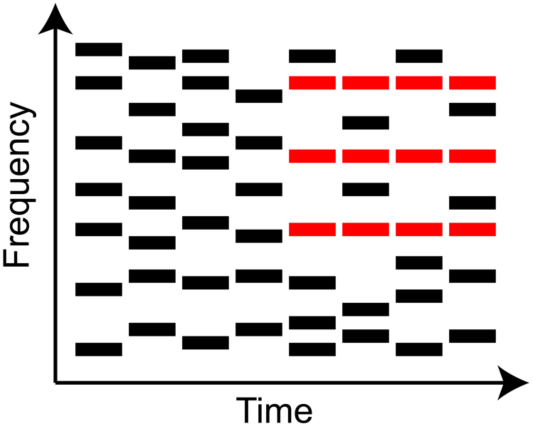
A schematic spectrogram of SFG stimulus.

All stimuli were created using MATLAB software (The Mathworks) at a sampling rate of 44.1 kHz and 16-bit resolution.

#### SFG: task

The SFG task was implemented in custom-written Matlab scripts (The Mathworks) using the Psychtoolbox 3 toolbox ((Brainard, 1997); (Pelli, 1997)). The SFG was conducted in an acoustically-treated, electrically-shielded booth with a single loudspeaker (model LOFT40, JBL) positioned at a 0° azimuth angle at a distance of 1.2 m. Visual stimuli were presented via a computer monitor located 0.5m in front of the subject at eye level. Sound levels were the same across subjects.

For each trial, participants saw a fixation cross on the computer screen. r a response was given.

### Statistical analyses

We related each predictor to speech perception performance on AzBio. The final model is given in (1), in the syntax of the linear modeling (i.e., lm()) function in R.

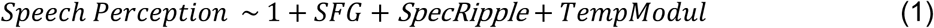

## Results

### Evaluation of independent variables in bivariate analyses

We started by evaluating the correlations among all the independent variables: to check for co-linearity prior to multiple linear regression analysis. No significant correlation was found between any predictor variables. The relationship between the predictor variables is shown in Figure 2 as scatter plots.

**Figure 2.**
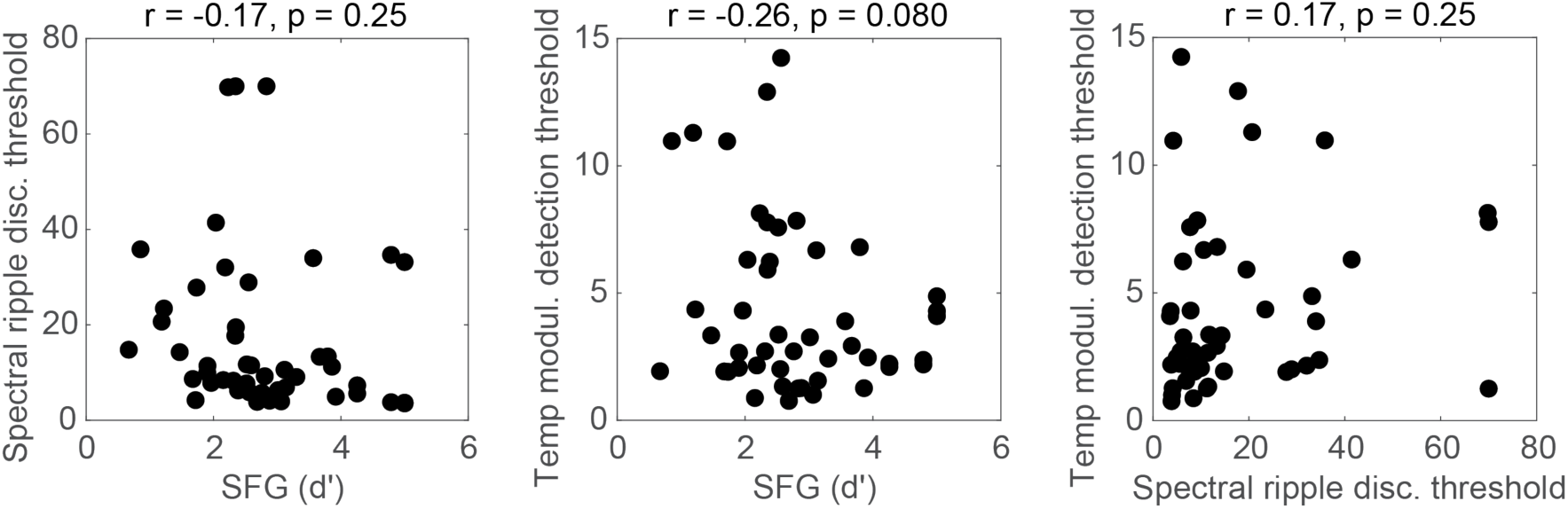
Results from predictor co-linearity analysis. No significant correlation is observed.

Next, we conducted exploratory bivariate analyses examining correlations between each independent variable and AzBio accuracy. All three predictors exhibited a statistically significant correlation with SiN ability. These are shown in Figure 3. In Figure 3, the spectral ripple discrimination and temporal modulation detection threshold values are in an arbitrary unit, measured as crossover during an adaptive procedure.

**Figure 3.**
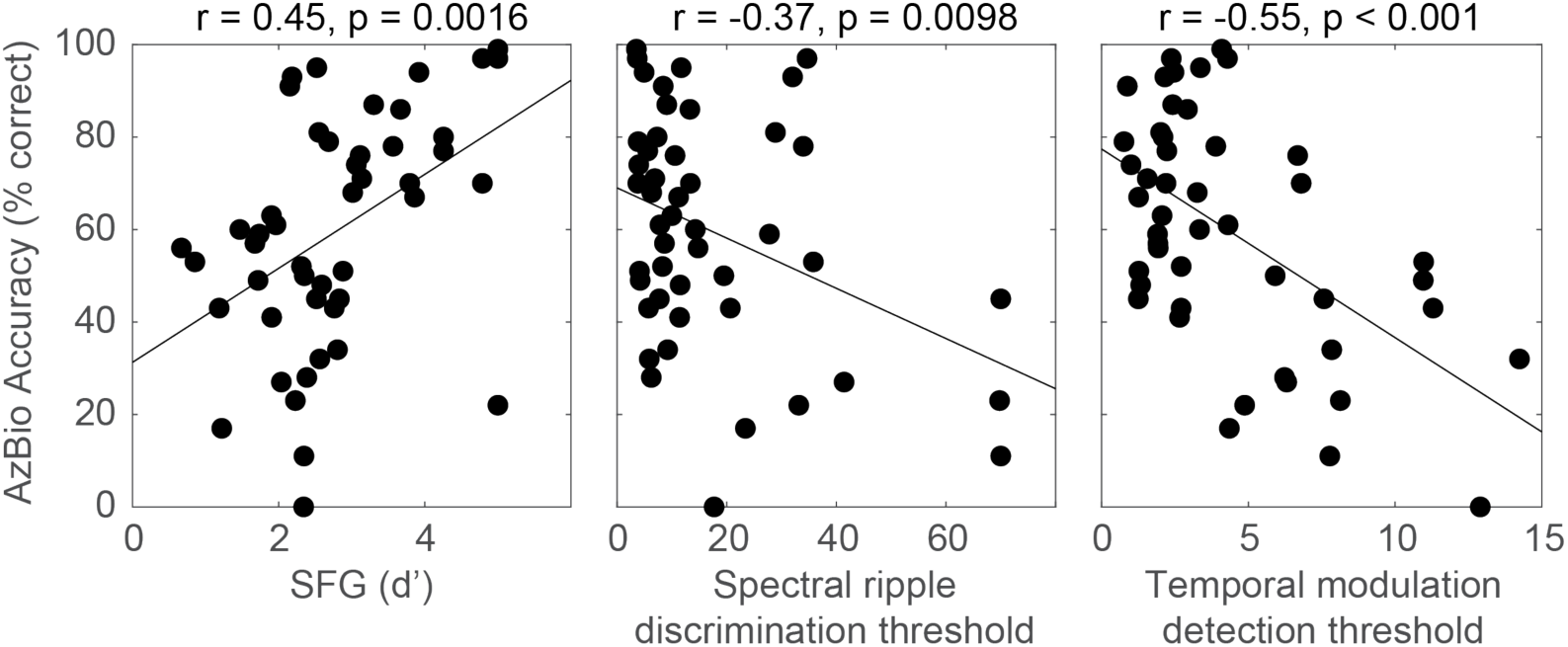
Results from bivariate correlation analyses. Spectral ripple discrimination and temporal modulation detection threshold values are in an arbitrary unit.

**Figure 4.**
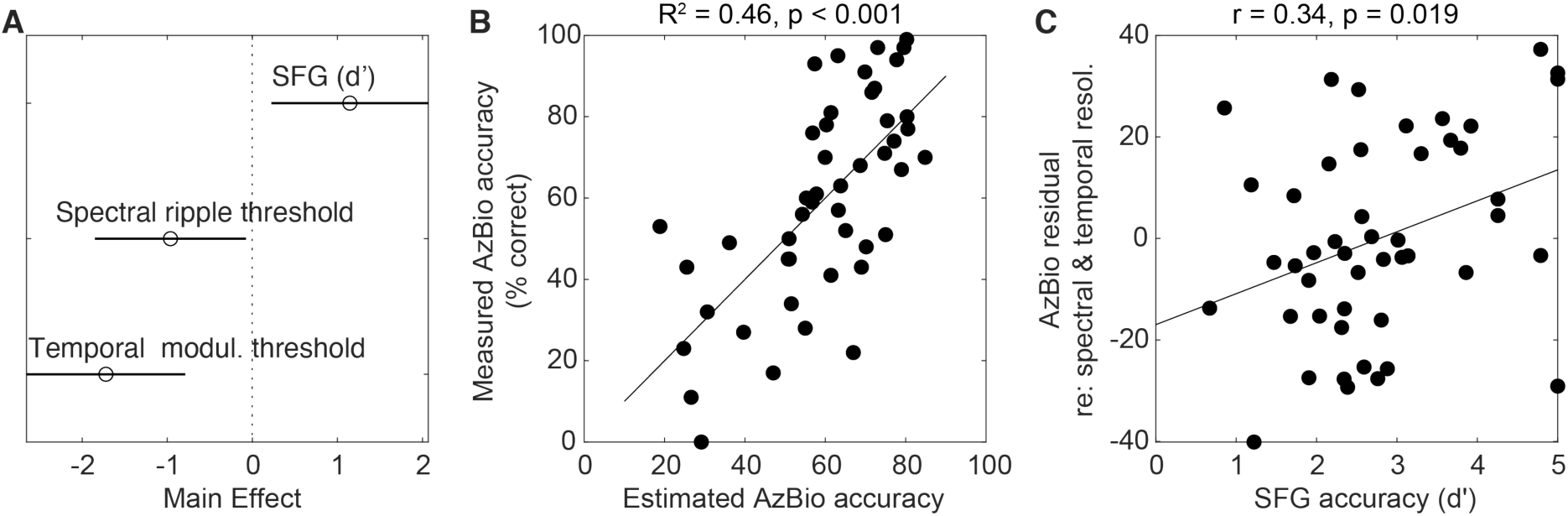
Results from multiple linear regression analysis. **A**. Main effects of predictor variables. **B**. Relationship between estimated AzBio accuracy (i.e., the model output) and measured AzBio accuracy (i.e., the dependent variable). **C**. Relationship between SFG accuracy and the residual of AzBio accuracy after regressing out the other two predictor variables (i.e., spectral and temporal resolution).

### Multiple linear regression

Following bivariate analyses, we conducted a multiple linear regression analysis to determine which of the independent variables predicted AzBio accuracy when accounting for all others (see Table 1 in its entirety). When adjusted for the number of independent variables, the model accounted for 46.3% of the variance in AzBio accuracy, *F*(3, 43) = 12.4, *p* <0.00001, Adjusted *R*^*2*^ = 0.426. All three predictors reached statistical significance.

**Table 1:** Results from multiple linear regression on SiN accuracy (N=47, R^2^=0.463):

| AzBio | $\beta$<br>(normalized) | SE | T(43) | p | Partial $\rho$ |
| --- | --- | --- | --- | --- | --- |
| SFG | 0.292 | 2.65 | 2.51 | 0.0160 | 0.357 |
| SpecRippleThres | -0.250 | 0.167 | -2.18 | 0.0345 | -0.316 |
| TempModulThres | -0.434 | 0.863 | -3.72 | <0.001 | -0.493 |

## Discussion

In this study, post-lingually deafened CI users performed a stochastic figure-ground (SFG) task in which listeners detect temporally coherent frequency components in the cloud of tone pips that rise at random times at random frequencies. The bivariate correlation between figure-detection performance (d-prime) and sentence-in noise performance (AzBio score) was high as r = 0.45 (p <0.005). The effect size is greater than that in normal hearing subjects reported in (Holmes & Griffiths, 2019). Moreover, multiple linear regression demonstrated a significant effect of figure detection (normalized beta coefficient = 0.29, p < 0.05) even after accounting for the fidelity of spectral and temporal encoding in the auditory periphery. The combined model explained 46% of the variance in SiN performance. This work has therefore established a relationship between a simple measure of the cross-frequency grouping of electrically coded signals, relevant to SiN ability.

This result suggests that auditory-cognitive mechanisms play a powerful explanatory in CI users’ outcomes in everyday communications. Adopting the SFG task in clinics may reveal a source of SiN difficulty in CI users that has not been recognized in the current clinical practice, which potentially opens up an opportunity for a novel aural rehabilitative strategy for CI users.

The figure detection ability during the SFG task is unlikely the only auditory cognitive mechanism that contributes to SiN performance. Although 47 was a relatively large sample size for a CI study, the number of predictor variables was limited to three to ensure a reasonable statistical power. A future study will consider more auditory-cognitive mechanisms (e.g., auditory working memory: (Akeroyd, 2008; Dryden et al., 2017; Kim et al., 2020)) as well as linguistic and general cognitive mechanisms.

## Data Availability

All data produced in the present study are available upon reasonable request to the authors.

## Acknowledgments

This work was supported by NIDCD P50 (DC000242 31) awarded to Gantz, Griffiths, and McMurray, Department of Defense Hearing Restoration and Rehabilitation Program grant awarded to Choi (W81XWH1910637), and DC008089 awarded to McMurray. The authors declare no competing financial interests.

## Notes

### Competing Interest Statement

The authors have declared no competing interest.

### Funding Statement

This study was supported by NIDCD P50 (DC000242 31) awarded to Gantz, Griffiths, and McMurray, Department of Defense Hearing Restoration and Rehabilitation Program grant awarded to Choi (W81XWH1910637), and DC008089 awarded to McMurray.

### Author Declarations

IRB of the Unversity of Iowa gave ethical approval for this work.

